# Validation testing to determine the effectiveness of lateral flow testing for asymptomatic SARS-CoV-2 detection in low prevalence settings

**DOI:** 10.1101/2020.12.01.20237784

**Authors:** Jack Ferguson, Steven Dunn, Angus Best, Jeremy Mirza, Benita Percival, Megan Mayhew, Oliver Megram, Fiona Ashford, Thomas White, Emma Moles-Garcia, Liam Crawford, Tim Plant, Andrew Bosworth, Michael Kidd, Alex Richter, Jonathan Deeks, Alan McNally

## Abstract

Lateral flow devices are quickly being implemented for use in large scale population surveillance programs for SARS-CoV-2 infection in the United Kingdom. These programs have been piloted in city wide screening in the city of Liverpool, and are now being rolled out to support care home visits and the return home of University students for the Christmas break. Here we present data on the performance of Lateral Flow devices to test almost 8,000 students at the University of Birmingham between December 2^nd^ and December 9^th^ 2020. The performance is validated against almost 800 samples using PCR performed in the University Pillar 2 testing lab, and theoretically validated on thousands of Pillar 2 PCR testing results performed on low-prevalence care home testing samples. Our data shows that Lateral Flow Devices do not detect infections presenting with PCR Ct values over 29-30, meaning that only 3.2% (95% CI 0.6% to 15.6%) of total cases in the student population were detected, but that as many of 85% of cases tested in the Pillar 2 PCR lab would have been detected theoretically

## Introduction

In November 2020 the United Kingdom government announced a plan to introduce mass scale population testing for SARS-CoV-2 infection using Lateral Flow Devices (LFD). Principal of these is an LFD manufactured and marketed by Innova Medical group, a subsidiary of Xiamen Biotime Biotechnology company. The LFD is a rapid lateral flow device based on colloidal gold immunochromatography designed to detect the presence of SARS-CoV-2 nucleocapsid antigens in nasopharyngeal swabs (1). The test can provide a result within 30 minutes allowing rapid testing on a mass scale.

The Innova LFD has very quickly been put into implementation by the Department for Health and Social Care (DHSC) and was employed in the city of Liverpool to deliver an ambitious mass-scale surveillance project of the city over a 2 week period (2). Data from the city council (3) shows that 71,684 LFD tests were performed alongside 51,855 gold-standard PCR tests (a total of 119,054 residents tested) with 439 people testing positive (0.37% positivity rate). The LFD tests are now being used in a pilot project to support people visiting relatives in Care Homes, and are being rolled out to support testing of University students before leaving campus to return to their family homes for the Christmas break.

In order to support this growing planned use of the Innova LFD test, the University of Oxford and Public Health England performed a series of validation trials of the LFD, benchmarking their performance against RT-PCR using swabs from a number of research trials in the United Kingdom (4). These included comparative testing on samples taken for the FALCON study evaluating diagnostic platforms, and bespoke trials including PHE, hospital, military staff and schools. The key headline findings of the validation report were that the LFD had a Limit of Detection (LoD) of around 100 plaque forming units/ml or 100,000 RNA copies/ml. In the report it is not made clear which RT-PCR assay is used in the comparison, but the Ct value given of 25.5 equating to 100 pfu/ml suggests it is not the ThermoFisher Covid-19 TaqPath assay (5) employed in the majority of Pillar 2 testing labs in the United Kingdom. As such the validation report may not fully indicate the potential of the performance of the LFD against the vast majority of Covid PCR testing done in the UK through Pillar 2

## Methods

### Lateral Flow device testing of students

As part of the national plan to test students for SARS-CoV-2 before the 2020 Christmas break, the DHSC provided University of Birmingham with Innova Lateral Flow Devices to test 15,000 students. Using a University owned booking system a total of 7,189 students were tested between December 2^nd^ and December 9^th^. Students were provided with a sterile nasopharyngeal swab and under supervision from a trained member of the University testing team, swabbed both tonsils and a single nasal cavity. The swab was passed through an opening in a plastic screen to a designated test area, where it was immediately processed according to the Innova protocol. Tests were performed by trained members of the University testing team drawn from post-graduate and final year undergraduate students in the College of Medical and Dental Science in the University, supervised by highly experienced post-doctoral researchers. In total a team, of 18 test operatives oversaw 36 testing booths, with a student attending a booth every ten minutes. A further 7 staff acted as results recorders logging the test results via a barcode through a DHSC provided mobile phone device and result recording app.

### Validation of Lateral Flow Device test results by PCR testing

University of Birmingham is home to a national pillar 2 testing laboratory, termed Turnkey lab, which conducts SARS-CoV-2 PCR diagnostics on behalf of DHSC (6). The laboratory uses the ThermoFisher Covid-19 taqPath assay used routinely in the Lighthouse laboratory testing network and tests a range of samples from mobile and stationary test sites (6). On each day of testing 90 residual Lateral Flow device test samples (saline solution in which the nasopharyngeal swab is resuspended to perform the test) were selected for confirmatory PCR testing. All positive samples were chosen for confirmatory PCR and the remainder were randomly selected samples. All samples were completely anonymous to the testing team with no identifying labels and were arbitrarily numbered from 1-90 each day. Sterile water was added to the samples to bring them to the 500 microlitres required for automated RNA extraction, and tested according to Pillar 2 laboratory protocol (6).

### Statistical analysis of PCR validation

The efficient stratified study design involved verification of all Innova test positives with RT-PCR, and a random sample of 720 of the 7187 Innova test negatives. Estimates of sensitivity and prevalence with 95% confidence intervals were obtained using maximum likelihood inverse probability weighted logistic regression to account for the sampling design, with conversion of the estimated odds to probabilities. Weights of 1 for Innova test positives, and 9.98 (7187/720) for Innova test negatives were used. Expected numbers of cases were computed from the prevalence estimate. The estimate of specificity was obtained without weighting as no Innova test positives were observed in those with negative PCR. Exact binomial methods were used to compute confidence intervals for test yield and specificity.

### Theoretical validation of LFD performance against Pillar 2 PCR test data

As part of pillar 2 testing our Turnkey laboratory also conducts PCR testing as part of the national Care Home surveillance plan implemented by DHSC to test all care home staff and residents to assist in control of Covid-19 transmission in UK care homes (7). Between October 25^th^ and November 5^th^ the Birmingham Turnkey laboratory processed a total of 19,176 PCR tests on home test and care home samples from across the United Kingdom. Of these 641 samples tested positive for SARS-CoV-2 using the cut off of two of three gene targets amplifying at a Ct value of 35 or under (6). This gives a positivity rate of 3.3%, around the rate that might be reasonably be expected in a large random surveillance of the UK population at that moment in time.

### Validation of Pillar 2 PCR positive samples on the Lateral Flow Device

We randomly selected ten anonymous samples from the Turnkey laboratory returning PCR positive results, encompassing samples positive at Ct values either side of the 29-30 detection cutoff. Two drops of sample were added using the sample tube provided in the Innova test kit, and results measured after 25 minutes. Additionally we tested 36 samples containing the very recently described new variant of SARS-CoV-2 identified in South East England, in which the S gene product does not amplify in our Thermo Taq-path assay.

## Results

### Lateral flow testing of University of Birmingham students

A total of 7,189 students voluntarily attended the asymptomatic student testing centre between December 2^nd^ and December 9^th^. Students were refused a test if they had any symptoms of COVID-19 and were referred to a local test site for PCR testing. Results of four samples were void, and two samples tested positive for SARS-CoV-2 by lateral flow, a prevalence of 0.03% (95% CI 0.02% to 0.10%) in the students volunteering for a test (Table 1).

**Table 1:** Table of results for Lateral Flow Device testing of University of Birmingham students and confirmatory PCR testing of approximately ten percent of samples.

|  | LFD tests |  | PCR Validation |  |
| --- | --- | --- | --- | --- |
| Day | Positive | Negative | Positive | Negative |
| 02/12/2020 | 0 | 630 | 0 | 90 |
| 03/12/2020 | 2 | 589 | 2 | 89 |
| 04/12/2020 | 0 | 1102 | 1 | 88 |
| 05/12/2020 | 0 | 860 | 1 | 89 |
| 06/12/2020 | 0 | 610 | 0 | 90 |
| 07/12/2020 | 0 | 813 | 2 | 88 |
| 08/12/2020 | 0 | 1259 | 2 | 88 |
| 09/12/2020 | 0 | 1320 | 0 | 90 |
| **Totals** | **2** | **7183** | **8** | **712** |

### Lateral flow results validation by Pillar 2 PCR

The two samples positive by Lateral Flow, and another 710 randomly selected negative samples were transported to the University Turnkey laboratory for PCR testing (9.9% of sample total). Of the 712 samples tested by PCR, 8 were positive: The two positive by Lateral Flow and a further 6 samples negative on the Lateral Flow Device (table 1).

Our PCR validation data suggests a true prevalence rate in the student population tested of 0.86% (95% CI 0.40% to 1.86%). The overall sensitivity of the test in the tested student population was observed to be 3.23% (95% CI 0.60% to 15.59%). We estimate that there would have been 62 cases in the 7185 students, of which 60 were missed. There were no false positive results, observed specificity was 100% (95% CI: 99.48% to 100.00%)

We further investigated the PCR testing discrepancy by extracting the Ct values for the amplification curves for the 8 PCR positive samples (Table 2). Our data shows that the six samples testing false Negative by Lateral Flow all had Ct values > 29, whilst the two true positive samples had Ct values of 20 and 25. We collated the RT-PR raw data from three technical replicates of assays performed on the Qnostics SARS-CoV-2 analytical Q-panel – 01 and generated average Ct values for each of the known viral titres provided in the panel (Table 3). Using this data we determined that at 100 viral copies per ml (the designated LoD for the Innova LFD – 3) the equivalent Ct values for the pillar 2 PCR assay would be a Ct of 30.8 based on the N gene target, roughly in line with our PCR validation data.

**Table 2:** Pillar 2 PCR Ct values of confirmatory samples positive for SARS-CoV-2. The samples which tested positive on Lateral Flow device are in grey columns.

| Well Number | ORF1ab Ct | N gene Ct | S gene Ct |
| --- | --- | --- | --- |
| 38 | 32.501118 | 33.89259 | 34.815563 |
| 1 | 25.160471 | 25.678833 | 25.034386 |
| 57 | 21.319279 | 22.638311 | 20.582413 |
| 36 | 28.538937 | 29.359957 | 29.123411 |
| 38 |  | 32.216614 | 33.92468 |
| 42 | 27.669174 | 29.401642 | 27.895124 |
| 14 | 30.770878 | 31.718863 | 31.998856 |
| 34 | 30.917858 | 31.794565 | 31.35588 |

**Table 3:** Analytical sensitivity and specificity of the Birmingham Turnkey lab RT-PCR pipeline. This was assessed against the commercial Qnostics SARS-CoV-2 analytical Q-panel – 01. Ct values are a median of 5 independent technical replicate, and figures in parentheses indicate the percentage of replicates returning a PCR positive for that given gene target (Ct < 35).

| QNostic Sample ID | Copies/ml | Log10 Copies | Orf1ab | S Gene | N Gene |
| --- | --- | --- | --- | --- | --- |
| SCV2AQP01-S01 | 1000000 | 6 | 15.3 (100%) | 16.5 (100%) | 15.8 (100%) |
| SCV2AQP01-S02 | 100000 | 5 | 18.3 (100%) | 20.5 (100%) | 17.4 (100%) |
| SCV2AQP01-S03 | 10000 | 4 | 22.8 (100%) | 24.9 (100%) | 23.6 (100%) |
| SCV2AQP01-S04 | 5000 | 3.7 | 19.5 (100%) | 24.8 (100%) | 24.1 (100%) |
| SCV2AQP01-S05 | 1000 | 3 | 25.9 (100%) | 28.6 (100%) | 26.6 (100%) |
| SCV2AQP01-S06 | 500 | 2.7 | 25.8 (100%) | 29.1 (100%) | 25.8 (100%) |
| SCV2AQP01-S07 | 100 | 2 | 27.7 (33%) | 30.3 (66%) | 30.8 (100%) |
| SCV2AQP01-S08 | 50 | 1.7 | 29.3 (17%) | 31.1 (27%) | 29.1 (55%) |
| SCV2AQP01-S09 | Negative | ---- | Negative | Negative | Negative |

### Sensitivity of Lateral Flow device in the student population by Ct value

From our data the Lateral Flow Device test yield is 2.8 per 10,000 (0.3 to 10 per 10,000). The sensitivity of the Lateral Flow device in the tested population differs greatly dependent on the viral titre of the person tested. At a PCR Ct value < 29 the sensitivity was 100% (95% CI 15.8 to 100). However, at a Ct <= 29 this dropped to 9.1% (1.03 to 49.1), and at Ct < 33 dropped again to 5.01% (0.78 to 32.14).

### Extrapolation of Pillar 2 PCR data to theoretically evaluate Lateral Flow device performance

We collated the raw RT-PCR data for all 641 of our positive samples as of November 5^th^ and ranked them according the N gene Ct value (Supplementary table). We then plotted the distribution of Ct values for our 641 positive samples (Fig 1). Using the LoD of 100 pfu/ml, we determined that this would correlate with an N gene Ct value of 30.8 plus one other gene target amplifying at a Ct < 35. By applying this theoretical level of performance to the LFD we determine that 99 of our positive samples would not be able to be detected by the Innova LFD given that the Ct value of N gene is above 30.8. This equates to 15.44% of our true positive RT-PCR samples being missed by the LFD. This means that the theoretically the Innova LFD when compared to Pillar 2 samples from low-prevalence, asymptomatic population screening similar to student and care home surveillance, would successfully detect 84.56% of all infections.

**Figure 1:**
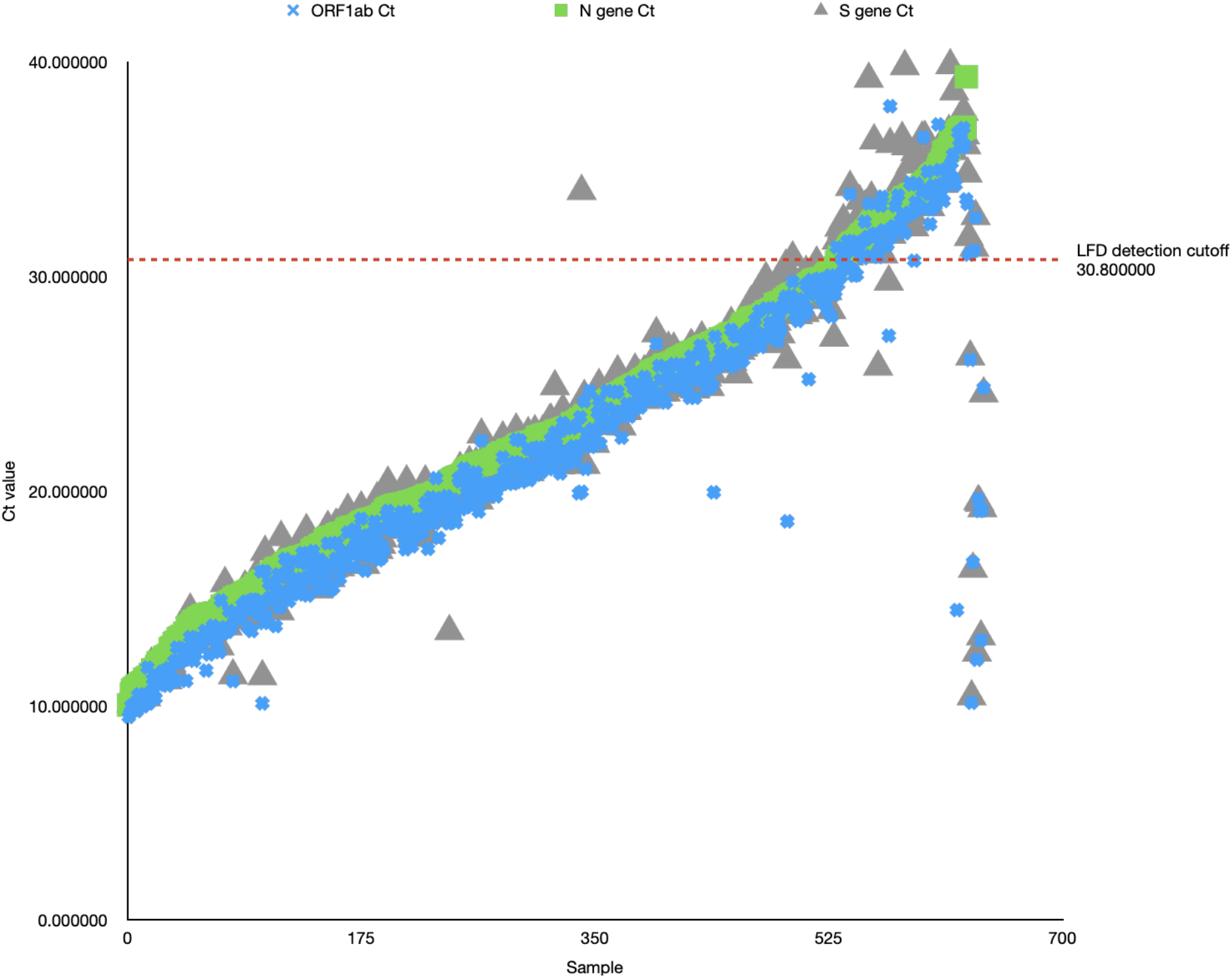
Graph plotting raw Ct values (Y-axis) for all 641 positive samples in the Birmingham Turnkey lab (Y-axis). Ct values for each of the targets (Orf1, N, S) are plotted, with a sample only called positive if at least 2 of the three targets have a Ct < 35. The red line indicates the N gene Ct value equating to 100 viral copies/ml, the previously determined LoD for the Innova LFD.

## Conclusions

Our data shows that the Innova Lateral Flow device can successfully detect SARS-CoV-2 infection in people with a viral titre above approximately 100 viral copies/ml. However it is incapable of detecting infection at comparable PCR Ct values of 30 and over. These levels of infection are indicative of very early or very late stages of infection, and as such we would strongly recommend that Lateral Flow Device testing is used to screen people at very regular frequency, and that a negative result should not be used to determine that someone is free from SARS-CoV-2 infection.

## Data Availability

All data is available in the supplementary file

## Notes

### Competing Interest Statement

The authors have declared no competing interest.

### Funding Statement

The testing work in this project is funded by the UK Department of Health and Social Care

### Author Declarations

The use of sample result data, and of anonymised waste samples from student testing programs in this study was reviewed by the Ethics review committee of the College of Medical and Dental Sciences at the University of Birmingham. A waiver was granted allowing samples to be used under ethics gained to aid assay development (NRES Committee West Midlands - South Birmingham 2002/201 Amendment Number 4, 24 April 2013)

### Summary of Updates

We have updated our manuscript with the results of almost 8000 LFD tests on University students

